# Microplanning improves stakeholders’ perceived capacity and engagement to implement lymphatic filariasis mass drug administration

**DOI:** 10.1101/2024.03.27.24304948

**Authors:** Caitlin M. Worrell, Tara A. Brant, Alain Javel, Eurica Denis, Carl Fayette, Franck Monestime, Ellen Knowles, Cudjoe Bennett, Jürg Utzinger, Peter Odermatt, Jean-Frantz Lemoine

**Affiliations:** U.S. Centers for Disease Control and Prevention, Atlanta, Georgia, USA; Swiss Tropical and Public Health Institute, Allschwil, Switzerland; University of Basel, Basel, Switzerland; IMA World Health, Port-au-Prince, Haiti; IMA World Health, Washington, DC, USA; National Program to Eliminate Lymphatic Filariasis, Ministry of Public Health and Population, Port-au-Prince, Haiti

## Abstract

**Background:** Achieving adequate mass drug administration (MDA) coverage for lymphatic filariasis is challenging. We sought to improve stakeholder engagement in MDA planning and improve subsequent MDA coverage through a series of microplanning workshops.

**Methodology/Principal Findings:** Prior to the 2018 MDA, Haiti’s Ministry of Public Health and Population (MSPP) and partners conducted 10 stakeholder microplanning workshops in metropolitan Port-au-Prince. The objectives of the workshops were to identify and address gaps in geographic coverage of supervision areas (SAs); review past MDA performance and propose strategies to improve access to MDA; and review roles and responsibilities of MDA personnel, through increased stakeholder engagement. Retrospective pre-testing was used to assess the effectiveness of the workshops. Participants used a 5-point scale to rank their understanding of past performance, SA boundaries, roles and responsibilities, and their perceived engagement by MSPP. Participants simultaneously ranked their previous year’s attitudes and their attitudes following the 2-day microplanning workshop. Changes in pre- and post-scores were analyzed using Wilcoxon-signed rank tests. A total of 356 stakeholders across five communes participated in the workshops. Participants conducted various planning activities including revising SA boundaries to ensure full geographic reach of MDA, proposing or validating social mobilization strategies, and proposing other MDA improvements. Compared with previous year rankings, the workshops increased participant understanding of past performance by 1.34 points (standard deviation [SD]=1.05, p<0.001); SA boundaries by 1.14 points (SD=1.30; p<0.001); their roles and responsibilities by 0.71 points (SD=0.95, p-<0.001); and sense of engagement by 1.03 points (SD=1.08, p<0.001). Additionally, drug coverage increased in all five communes during the 2018 MDA.

**Conclusions/Significance:** Participatory stakeholder workshops during MDA planning can increase self-reported engagement of key personnel and may improve personnel performance and contribute to achievement of drug coverage targets. Microplanning success was supported by MDA results, with all communes achieving preset MDA coverage targets.

**Author summary:** Lymphatic filariasis is a neglected tropical disease that can be eliminated by treating entire at-risk communities with safe and efficacious medicines, a strategy known as mass drug administration (MDA). MDA campaigns require intense planning to ensure that every eligible person within the community can receive the medicines if they desire. We aimed to improve the campaign by better involving key MDA stakeholders such as volunteers and other important community members in the planning process, through microplanning. The participants suggested many strategies to help the campaign reach more community members, including better ways to prepare and inform that community that the campaign is happening. We invited the microplanning participants to give feedback on how this new strategy worked compared with their experiences during past campaigns. Participants reported that they felt better engaged by health authorities, and in particular, that they had more information about the results of past campaigns, where they should be distributing medicines, and their specific tasks and responsibilities during the campaign. We found that more people received medicines during the campaign that followed the microplanning workshops compared with the previous campaigns. We conclude that microplanning helped to increase the number of people who received MDA medicines.

## Introduction

Lymphatic filariasis (LF) programs in some countries have met global program targets for elimination. However, many programs continue to face challenges in achieving high levels of community participation in mass drug administration (MDA) [1]. Triple drug regimen MDA [2] may accelerate national and global progress toward LF elimination [3], but the benefits of such novel approaches can only be realized if a high proportion of at-risk community members have access to the intervention and are receptive to taking the drugs [3]. Numerous factors can impact community drug coverage and compliance [4, 5], and the factors that influence an individual’s willingness and ability to participate in MDA are multi-factorial and subject to change through multiple rounds of MDA [5]. These factors are often related to the broader social-ecological context, as well as provider or recipient community characteristics.

While many studies have highlighted the recipient community characteristics that drive non-adherence to MDA [6–8], increasing focus has been placed on understanding and addressing provider-related issues that are important drivers of drug uptake [9]. In Tanzania, for example, the majority of community members who did not receive the medication reported program-related issues, such as community drug distributors (CDDs) not visiting all eligible households, inopportune timing of the MDA, and not knowing that MDA was occurring [10]. Other reported program issues associated with poor MDA compliance include drug shortages, insufficient time and human resources, inadequate training of drug distributors, inappropriate selection of drug distributors, poor adverse event management, and limited community engagement or coordination in MDA planning [5]. Failure to adequately engage members of the health system and the community during MDA planning can adversely impact the quality of MDA implementation and, therefore, lead to low levels of community participation [11, 12]. Conversely, strong community participation in the planning and implementation of MDAs has been seen as key to the success of some campaigns, particularly in urban areas [13–15]. In India, the involvement of mid-level health authorities, especially at the district level, was identified as a key element in carrying out a successful MDA [6].

Microplanning is an approach that may improve access to and acceptability of MDA programs by addressing the various issues faced by providers and recipients [16]. While the specific implementation varies by location, microplanning generally encompasses a ‘bottom-up’ approach that engages local stakeholders in health program planning by leveraging local data and knowledge to both identify site-specific problems and design solutions [17]. This iterative and multi-stage process engages local representatives to define the target populations in a delineated area that are eligible for an intervention [18]. During this process, stakeholders define the location of the target population and the catchment areas for each program delivery actor. Stakeholders design service delivery strategies to reach targeted sub-populations, often with an emphasis on hard-to-reach populations (e.g., mobile and remote populations). They create a realistic operational plan based on local circumstance and available resources, as well as determine the material, financial, and human resources needed to accomplish program targets. Microplans are typically gathered and compiled across various administrative levels and, ideally, inform the allocation of program resources.

Microplanning has been shown to improve public health program coverage [19], and is a key component of the Reaching Every District strategy [18, 20] for improving immunization systems and vaccination coverage [19, 21, 22]. Outside immunization programs, microplanning has been used in planning and delivering other health interventions, including HIV care and prevention [23, 24], malaria prevention [25, 26], and reproductive and child health programs [27, 28]. Neglected tropical disease (NTD) programs such as LF MDA have applied microplanning less frequently than other programs due to financial constraints. However, as national NTD programs approach LF elimination targets and identify areas of LF foci of transmission that appear to be particularly intractable, new tools and strategies are needed. Increasingly, digital tools are being used during microplanning for health program planning and implementation [25, 29] as they have been shown to be a cost-effective strategy [25, 30] that can increase program coverage [23, 31], identify mobile and displaced populations [21, 29, 31], and promote an efficient allocation of resources [32].

Haiti has been implementing large scale MDA campaigns for LF since 2000 and reached full scale in 2012 once MDA began in the metropolitan region of Port-au-Prince [33]. Despite most of the communes in Port-au-Prince achieving globally recommended targets of at least 65% population coverage of MDA for LF during initial rounds [34], the Port-au-Prince communes struggled to achieve coverage targets during subsequent rounds of MDA [35]. In 2016, sentinel site assessments identified that five of six Port-au-Prince communes required additional rounds of MDA that achieved coverage targets prior to qualifying to undergo the transmission assessment survey (TAS). After the 2017 MDA, in which coverage targets were again not met, the Ministry of Public Health and Population (MSPP) convened a meeting with implementing partners in order to create a plan for improving coverage in subsequent MDAs. During these meetings, the partners identified a series of strategies for improving coverage based on literature of best MDA practices in urban areas and lessons learned by MSPP staff and partners during years of MDA implementation. Microplanning was one of several proposed activities.

The study reported here details the results of a microplanning activity aimed to improve 2018 MDA coverage in the five communes of Port-au-Prince that had experienced declining MDA coverage. Our specific objectives were to evaluate the effect of using established microplanning techniques on (i) identifying program gaps and key solutions; (ii) the perceived performance and engagement of key stakeholders (both community leaders and program staff) who attended a microplanning workshop; and (iii) MDA coverage in the subsequent 2018 MDAs conducted in five communes of Port-au-Prince.

## Methods

### Ethics statement

This project took place as an evaluation of a routine LF elimination program and was considered by Haiti’s MSPP to be a program evaluation. The project is covered by a non-research determination granted by the Center for Global Health (CGH) Human Subjects Protection Office at the U.S. Centers for Disease Control and Prevention (CDC) in Atlanta, Georgia, USA.

### Project area

This evaluation took place from January through March 2018 in five of the six communes (i.e., Carrefour, Cité-Soleil, Delmas, Port-au-Prince, and Tabarre) comprising metropolitan Port-au-Prince. This area, which represents a population of approximately 2.3 million people, was the last area in Haiti to initiate LF MDA in 2012 [33].

In Port-au-Prince, MDA is delivered through distribution posts located in the community and at schools. One post, staffed by three CDDs, is allocated to treat approximately 1,000 people over a 4-day MDA. Community promoters (CPs) manage approximately three health posts and are in turn managed by community leaders (CLs). CLs, with assistance from CPs, are responsible for conducting social mobilization activities within a supervision area (SA) in the months prior to the MDA, selecting and supervising distribution sites, and training CPs and CDDs.

### Overall design

To improve stakeholder engagement and MDA implementation, we conducted a 2-phase microplanning exercise in the five target communes. In phase one, LF program staff, in collaboration with CLs, conducted an inventory of the 2017 MDA distribution posts using ODK Collect (Open Data Kit Inc., v2.0) [36] loaded onto mobile devices. Historic distribution post sites were overlaid onto a Google satellite base layer (Google Inc, n.d.), using open-source QGIS (Open Source Geospatial Foundation Project, v2.14.21). To identify possible access gaps, we applied a 300 m buffer around each distribution post to identify areas within approximately a 500 m maximum walking distance. These maps were prepared to inform the discussion during microplanning workshops in phase two.

During phase two, all CLs as well as a selection of CPs, and other community stakeholders were invited to participate in microplanning workshops. In close consultation with communal and departmental focal points from MSPP, the program staff identified key community stakeholders who would be critical for community- and school-based with distribution. These individuals included school directors, Ministry of Education inspectors at the district and sub-district level. For community-based MDA this included representatives from local authorities such as mayors and CAESECs, church-leaders and pastors, voodoo temple, and local youth associations. The microplanning workshops included several key activities. Initially, the project staff provided an overview of the LF elimination program objectives, reviewed the historical results from the 2012 to 2017 MDAs, summarized the results of several key MDA evaluations, and engaged the participants in a discussion of the challenges to and opportunities for improving MDA coverage. To clarify stakeholder roles and responsibilities, program staff presented and validated the terms of references for key MDA staff (i.e., CLs, CPs, and CDDs), and drafted a document detailing expectations from opinion leaders and other stakeholders. Further, program staff presented several specific strategies that were proposed by implementing partners to improve the 2018 MDA. These strategies included but were not limited to extending the number of MDA days, modifying MDA distribution times, enhancing schools’ participation in MDA, improving visibility and credibility of CDDs, and expanding mop-up activities. Participants were invited to propose revisions including defining and validating key strategies for community drug distribution (e.g., optimal timing and location).

Finally, SA boundaries were delineated for each community leader in QGIS (Open Source Geospatial Foundation Project, v2.14.21). GPS coordinates of distribution posts collected during phase one were overlayed onto an Open Street Maps layer that included various landmarks (e.g., streets, places of business, etc.). Google Maps and satellite base layers (Google Inc., n.d.) were used as additional references for identifying landmarks and delimiting boundaries. Through an iterative process, microplanning participants used the 2017 distribution post maps as a basis to discuss and agree upon the supervision area boundaries for each CL, ensuring no overlapping or omission of areas within the targeted communes.

### Impact measures

We used retrospective pre-testing to evaluate the microplanning workshops against the stated objectives [37–39]. This method involves asking participants in a single data collection event to rate their attitudes before and after the workshop. Following the microplanning workshops, participants were invited to complete a questionnaire simultaneously rating their previous year’s perceptions and their perceptions following the 2-day microplanning workshop. This strategy aimed to reduce the confounding factor of response shift bias [40], a phenomena where the respondents’ internal frame of reference is altered due to the influence of the intervention itself. In this context, the participants perceptions of their previous knowledge and engagement may be altered by participating in the workshop, as has been seen in evaluations using traditional self-reported pre-post-test evaluation frameworks[41]. Participants were asked to evaluate four measures: (i) understanding your past performance; (ii) understanding the boundaries of your coverage zones or area of influence; (iii) understanding your roles and responsibilities in the MDA; and (iv) engagement by MSPP in the MDA planning process. Participant commune and role in the MDA were also recorded. Each measure was evaluated using a 5-point scale where 1=poor; 2=fair; 3=good; 4=very good; and 5=excellent; N/A=not applicable. We also assessed changes in coverage between the 2017 and 2018 MDAs as an indirect measure of impact. MDA coverage was calculated using the total number of MDA doses delivered by each commune’s MDA distribution teams divided by the estimated population of the commune.

### Statistical analysis

Data were compiled using spreadsheet software (Microsoft Excel 2010, Microsoft, Seattle, WA, USA) and data management and statistical analyses were completed using SAS v9.4 (SAS Institute, Cary, NC, USA). For pre-workshop and post-workshop summary analyses, all responses were considered, however results were filtered to include only complete pre-post pairs to measure changes as a result of the workshop. Within pair differences between the pre- and post-workshop ratings were analyzed using a Wilcoxon signed-rank test to determine whether microplanning had impacted perceived capacity to deliver quality MDA as well as key engagement metrics. Alluvial plots describing changes in participant ratings were created in R v1.1.463 (RStudio, Free Software Foundation Inc., Boston, MA, USA).

## Results

### Microplanning outputs

During phase one, project staff and CLs collected 8,034 GPS coordinates representing the 2017 distribution points. In advance of the microplanning workshop, the GPS coordinates were plotted in QGIS overlayed on satellite images of Port-au-Prince to create a detailed distribution post map for all five communes. Program staff used these distribution post inventory maps to assess coverage gaps that were presented to MDA stakeholders.

During phase two, 10 2-day microplanning workshops were held for 356 participants, including 73 CLs, 240 CPs, and 43 other key MDA stakeholders selected from the five targeted communes. Several activities were conducted as part of the workshops. First, LF program staff presented workshop participants with the maps showing the location of the 2017 distribution posts. Staff then worked with CLs to delineate the 2017 SA boundaries as they were understood by each CL. **Supplemental Figure 1** shows the resulting SA maps following the iterative process by which LF program staff and workshop participants modified the MDA maps to ensure no overlapping or omitted areas within the targeted communes. As part of this process, certain leaders’ distribution posts that fell outside their updated SA boundaries were flagged to be prioritized for relocation.

To support the optimal deployment of posts, each CL received a printed map following the workshop that contained detailed information about their supervision area, including roads and points of interest such as churches, businesses, and schools (**Figure 1**). Each CL’s supervision area was clearly outlined, and neighboring CLs were listed. Maps were printed in Haitian Creole to improve the ease of use by stakeholders and listed the CL’s name to foster a sense of ownership over the activities in that area.

**Fig. 1.**
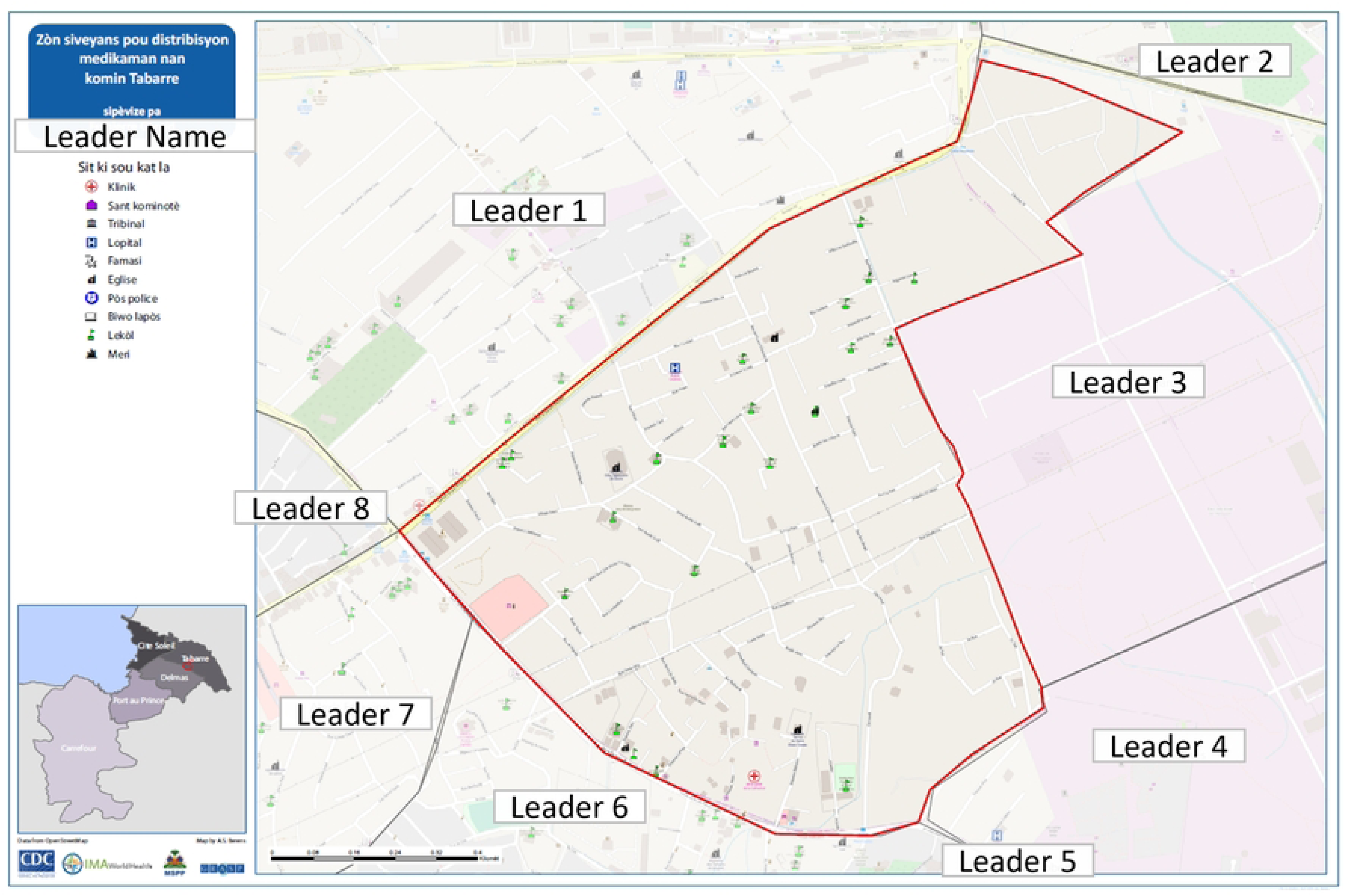
Illustrative supervision area (SA) maps, 2018 mass drug administration (MDA) in Port-au-Prince, Haiti. Illustrative supervision area (SA) map displaying the SA for a community leader in the commune of Tabarre for the 2018 mass drug administration for lymphatic filariasis (in Haitian Creole). The map includes the boundaries of the SA (in red) including infrastructure and key points of interest obtained from OpenStreetMaps. Additionally, it shows the community leaders working in adjacent SAs. For privacy reasons, this figure has been lightly edited to anonymize the names of community leaders.

**Figure 1** shows an illustrative CL’s supervision map.

Another key activity included reviewing past MDA performance, identifying local challenges to achieving high coverage, and proposing updated strategies to improve program coverage. As part of this process, participants discussed and finalized distribution strategy modifications including some modifications proposed by LF program staff, as well as drafts of new social mobilization strategies and tools that were developed by LF program staff and partners. **Table 1** describes a selection of key discussion points and associated solutions based on these discussions. Participants identified key actions to improve partnerships, planning and logistics associated with MDA planning; strategies to improve support for CLs, CPs, and CDDs; new social mobilization strategies; as well as modifications to improve distribution strategies.

**Table 1:** Selected feedback and outcomes from microplanning workshops conducted in Port-au Prince, Haiti in 2018.

| **Theme** | **Identified issues / key suggestions** | **Decisions or actions taken<sup>1</sup>** |
| --- | --- | --- |
| *Partnerships, planning, and logistics* | Some school authorities refuse to participate in MDA activities due to lack of timely official correspondence from Ministry officials. | Departmental authorities will prepare official correspondence detailing the specifics of the MDA distribution. This correspondence will be available to MDA staff within one week of the microplanning workshops. |
|  | Local authorities should accompany leaders during mobilization and promotion events at schools, churches, and other settings. | Local authorities are involved in the microplanning workshops to ensure that they are informed of the MDA activities and may coordinate with MDA staff to mobilize and sensitize their populations. |
|  | Boundaries of supervision areas are poorly understood by leaders and other authorities. | Supervision area boundaries are defined during the microplanning workshops. Detailed maps of the newly defined supervision areas, including boundaries, infrastructures, and key points of interest, will be distributed to leaders to facilitate efficient MDA planning. |
| *Supporting leaders, promoters, and CDDs* | Working conditions should be improved for the leaders to achieve better results. | Leaders and promoters will be provided official badges to facilitate their identification and perceived legitimacy, particularly at institutions such as schools. |
|  | Existing social mobilization tools are outdated and insufficient for adequate social mobilization of the community. | New social mobilization tools will be created as part of broader MDA strengthening activities. Training sessions will be held for the main MDA actors orienting them on the use of new social mobilization tools. |
| *Social mobilization strategies* | The program should create testimonials or documentaries featuring persons affected by LF disease. | Radio spots will be released promoting the MDA that include testimonials of persons affected by LF disease. |
|  | Local celebrities and community leaders should be better engaged in social mobilization campaigns. | A local comedian will be engaged to create social mobilization messages that will be disseminated on social media platforms.<sup>2</sup> |
|  | High level officials and medical professionals should take medications in public to improve public confidence in the reliability of the medicines. | A public launch of the MDA will be held and local authorities and partners will swallow the MDA medicines in public. |
|  | The program should publicize the location of sites that can manage adverse events during the MDA. | The program will make adverse event management information available through a toll-free hotline. |
| *Distribution strategies* | Schools are a key population that should be emphasized during MDA, however limited school information is available to staff, including lists and enrollment numbers of schools in the treatment zone. | Schools will be prioritized during MDA. Data on school population will be collected in collaboration with the Ministry of National Education and Vocational Training (MENFP). |
|  | MDA distribution hours should be shifted to include | Several communes will shift daily distribution times from 09:00 – 17:00 to |
|  | early evening hours to capture populations who are working or unavailable during working hours. | 10:00 – 18:00 hours; however, other communes will maintain the original distribution hours due to security concerns. |
|  | Leaders and drug distributors have difficulty reaching populations who live/work in restricted access zones (e.g., industrial areas or gated communities). | For leaders affected in restricted access zones, a specific distribution strategy will be developed with the list of key partners in support. |
|  | Some zones have insufficient personnel to adequately serve the targeted populations. | Two additional promoters will be assigned to support MDA activities in the Grand Ravine and Tibwa area of Port-au-Prince commune. |
|  | Upon harmonization of supervision area boundaries, stakeholders recognized that several MDA distribution posts were located in neighboring supervision areas or communes. | Several leaders' posts were identified for re-location to access. A Cité-Soleil leader will supervise posts in bordering Port-au-Prince, due to hesitation of Port-au-Prince leaders to work there due to insecurity. However, these posts will be counted as part of the Port-au-Prince coverage figures. |
|  | Several areas had insufficient distribution post coverage (e.g., mountainous areas within Carrefour commune). | Distribution posts will be reorganized to provide better coverage of the supervision areas, particularly in the mountainous areas of Carrefour commune. |
|  | Mop-up should be conducted after the initial MDA campaign to maximum coverage. | Both passive and active mop-up strategies will be implemented after the 5-day MDA campaign. |

<sup>1</sup> Some actions described in this table were devised by partners based on feedback from the microplanning process or other MDA improvement activities in response to stakeholder feedback and suggestions
<sup>2</sup> Link to social media https://www.instagram.com/p/BifOhCVBFfM/?utm_source=ig_web_copy_link

Several challenges were identified that were unable to be addressed fully by the LF program at the time of the activity (e.g., participants from multiple communes noted a need to improve working conditions for the leaders to achieve better results, noting a particular need for umbrellas at distribution posts and increasing MDA stipends). Participants from Carrefour noted that lack of health facilities in rural sections of communes pose a challenge for adverse events management. Participants from Cité-Soleil noted the need to combat LF at different stages in the transmission cycle, especially as effluence from factories creates sanitation concerns within the commune. Finally, the LF program staff presented and validated the roles and responsibilities of CLs, CPs, and CDDs.

### Respondent evaluation

Following the workshop, participants simultaneously evaluated their perceptions of their capacity and engagement prior to and following the microplanning workshops using a retrospective pre-test model. Evaluation questionnaires were completed by 339 (95.2%) workshop participants across all five targeted communes, including two (0.6%) national-level MSPP staff, two (0.6%) departmental-level MSPP staff, 68 (20.1%) community leaders, 227 (67.0%) community promoters, and 40 (11.8%) community members (**Table 2**). Participant counts were approximately proportional to the underlying commune population.

**Table 2:** Demographic characteristics of microplanning workshop participants (n=339)

| Characteristics | No. (%) of respondents |
| --- | --- |
| **Commune\*** |  |
| Carrefour | 72 (21.2%) |
| Cité-Soleil | 70 (20.7%) |
| Delmas | 73 (21.5%) |
| Port-au-Prince | 96 (28.3%) |
| Tabarre | 28 (8.3%) |
| **Role** |  |
| National MSPP | 2 (0.6%) |
| Departmental MSPP | 2 (0.6%) |
| Community leader | 68 (20.1%) |
| Community promoter | 227 (67.0%) |
| Community member | 40 (11.8%) |

\*Estimated population by commune in 2018: Carrefour=534,702; Cite-Soleil=277,180; Delmas=413,315; Port-au-Prince=1,032,409; Tabarre=136,234.

At baseline, understanding roles and responsibilities had the highest score 3.91 ± 0.91 (mean ± standard deviation, SD), followed by perceived engagement 3.41 ± 1.07, SAs 3.29 ± 1.12, and past performance 3.05 ± 1.00 (**Table 3**). The highest proportion of individuals providing a “poor” assessment was seen within respondents’ understanding of the boundaries of supervision areas (n=8, 21%), while the highest proportion of individuals reporting “excellent” was seen with understanding roles and responsibilities (n=73, 28%).

**Table 3:** Retrospective pre-testing results collected following microplanning workshops held in Port-au-Prince, Haiti in 2018.

|  | Pre-workshop |  | Post-workshop |  | Difference (post-pre) |  |  |
| --- | --- | --- | --- | --- | --- | --- | --- |
|  | n | Mean (SD) | n | Mean (SD) | n* | Mean (SD) | p-value |
| **Past performance** | 303 | 3.05 (1.00) | 302 | 4.37 (0.72) | 283 | 1.34 (1.05) | <0.001 |
| **Roles/responsibilities** | 277 | 3.91 (0.91) | 283 | 4.61 (0.60) | 262 | 0.71 (0.95) | <0.001 |
| **Supervision areas** | 277 | 3.29 (1.12) | 286 | 4.39 (0.82) | 266 | 1.14 (1.30) | <0.001 |
| **Perceived engagement** | 281 | 3.41 (1.07) | 286 | 4.45 (0.79) | 268 | 1.03 (1.08) | <0.001 |

\*Only individuals with complete pre- and post-workshop data were considered for a difference in pre- and post-workshop scores.

When assessing the participants’ scores after the microplanning session, the elements maintained the same rank order as in in the pre-workshop for the mean score for each metric, which were in descending order (post-test mean ± SD): roles and responsibilities (4.61 ± 0.72), perceived engagement (4.45 ± 0.79), SAs (4.61 ± 0.60), and past performance (4.37 ± 0.72).

**Figure 2** shows the evolution of pre-post responses for each respondent, among complete pairs.When comparing pre-and post-workshop scores, statistically significant improvements were seen in scores across all four evaluation metrics (p<0.001). The smallest improvement was seen among respondents’ understanding of their roles and responsibilities with 0.71 ± 0.95, while the greatest improvement was seen in understanding of past performance with a mean improvement of 1.34 ± 1.05 (**Table 3**).

**Fig 2.**
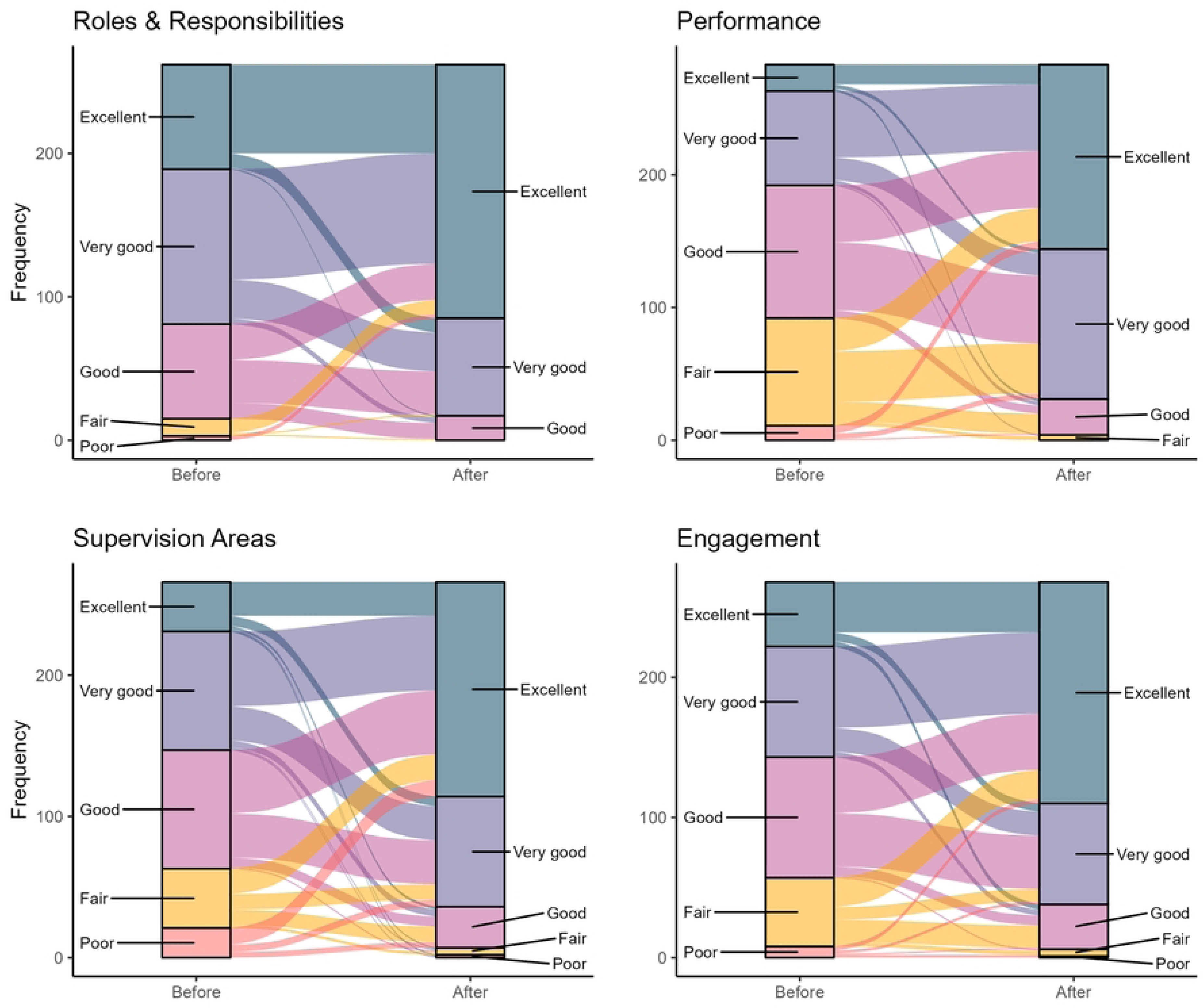
Changes in participant understanding and perceptions before and after microplanning workshops, Port-au-Prince, Haiti, 2018. The alluvial charts track the individual-level dynamics in ratings before and after the microplanning workshops for four indicators: (a) understanding of past performance; (b) understanding of SAs/zones of influence; (c) understand respective roles/responsibilities in the MDA; and (d) perception of engagement by MSPP in the MDA planning process. The left of each chart displays the distribution of participants’ ratings before the workshop, while the right indicates the participant responses from after the workshop. Participants ranked their perception on a 5-point scale where: 1=poor; 2=fair; 3=good; 4=very good; and 5=excellent. The width of the band corresponds to the relative weight of persons with a particular before-after combination. The color of the band corresponds to the score at the before timepoint, with red/orange colors representing lower before workshop responses.

The following alluvial charts show the individual-level dynamics in rating before and after the stakeholder workshop. On the left of each chart, you see the distribution of rating before the workshop with 1 representing “Poor” and 5 representing “Excellent,” while the numbers on the right of the chart indicate the responses after the workshop. The thickness of the band represents the number of people giving a particular response combination. The color of the band indicates the response at the time point before the workshop; with orange/red colors representing lower before workshop responses. For example, this band shows the 25 individuals who reported having a “Fair” understanding of their past performance prior to the workshop, but an “Excellent” understanding of past performance following the workshop.

### MDA coverage

Based on results of the microplanning meeting, the 2018 MDA campaigns in all five communes were extended from 4 to 5 days. MDAs were held from April 26 to June 4, 2018, sequentially in Tabarre, Cité-Soleil, Port-au-Prince, Carrefour, and Delmas. Reported MDA coverage increased in all five communes between 2017 and 2018 respectively with an increase from 50% to 120% in Tabarre, 50% to 93% in Cité-Soleil, 38% to 81% in Port-au-Prince, 37% to 72% in Carrefour, and 45% to 70% in Delmas.

## Discussion

### Microplanning outcomes

These evaluation results suggest that microplanning led to several important outcomes. First, the lack of clearly defined and communicated supervision area boundaries was a central challenge to ensuring broad access to the MDA in previous years. Using microplanning, program staff were able to collaboratively define SA boundaries with all key stakeholders, thus minimizing likelihood of missing underserved areas, efficiently allocating scare MDA resources, and avoiding duplication of efforts of a limited workforce across the Port-au-Prince region. With this knowledge, stakeholders were able to collaboratively realign SA boundaries and commit to deploying posts within their respective SAs. The use of digital mapping tools was critical for helping MDA stakeholders to visualize the targeted MDA area and harmonize boundaries of their SA, without which harmonizing SA borders would have been extremely challenging in this dense urban setting. Zonal maps provided needed documentation that allowed CLs and CPs to plan and execute activities in their assigned areas, as well as more effectively coordinate with staff in neighboring areas. The presence of numerous landmarks in urban settings and orienting microplanning participants to their individual zone helped with the uptake and use of the maps by CLs and CPs.

Second, the participatory workshops provided a forum for engaging various stakeholders in identifying key MDA challenges and collectively designing locally appropriate, feasible and acceptable solutions. For example, MDA staff from Port-au-Prince commune revealed that they did not cover certain areas within the commune due to safety concerns. One such area was an economically disadvantaged and underdeveloped area. In light of this, a CL from Cité-Soleil volunteered to cover these areas to ensure that no service gaps existed. Furthermore, volunteers from the Delmas commune expressed frustrations of being scheduled as the final commune to undergo MDA in the area as well as concern that CLs from neighboring communes placed posts within their commune boundaries. They feared that these factors artificially reduced their coverage figures, as members of the population were already treated by neighboring communes. These concerns were supported by coverage data showing progressively decreasing coverage figures as the MDA progressed sequentially across communes. The refinement of SA boundaries and creation of zonal maps reduced the risk of cross-commune post deployment.

Third, the microplanning activities provided previously unknown data and information that was helpful in supporting other MDA strengthening initiatives. For instance, the distribution post census allowed for the generation of unique identification codes for each distribution post. These codes were then used to track post-based distribution data through real-time data collection, which permitted MDA staff to track the daily progress of MDA in near real time and adapt MDA procedures to achieve adequate coverage.

Finally, one of the most successful outcomes of the microplanning was an increase in community engagement, program ownership, and motivation of MDA volunteers and stakeholders. By working with CLs and other stakeholders to review distribution post distribution and refine SA boundaries, program staff sense that stakeholders felt they were involved with the “highly technical work” of MDA planning, rather than passive actors. For example, several leaders took an initiative to conduct informal, uncompensated mop-up activities beyond their allocated MDA days. Increased motivation was corroborated by the results of the pre- and post-workshop surveys from participants attending the microplanning workshop. The survey data provided additional insights into the perceived utility of such participatory planning efforts from the perspective of MDA stakeholders. For example, although LF program staff perceived themselves to be generally successful in conveying the MDA roles and responsibilities to the various MDA actors, they reported further improvements after the microplanning exercise when roles and responsibilities were reviewed and validated collectively.

The workshop evaluation supported research by Wodnik *et al.* with CLs, CPs, and CDDs in Tabarre and Carrefour communes [35]. Their evaluation suggested that the LF program needed to improve communication and feedback of the outcomes of the MDA rounds with the stakeholders, as well as provide opportunities for collectively identifying and implementing strategies to improve MDA [35]. Further, their evaluation revealed that SAs were poorly understood by CLs, likely leading to missed populations during previous MDA activities. Finally, they suggested that strategies such as microplanning can be a successful approach for engaging MDA staff and can contribute to increased program ownership and improved MDA outcomes.

We believe the experience in Port-au-Prince highlights several considerations that programs seeking to increase MDA coverage though microplanning in other settings, particularly complex urban locations, should consider. The strategy employed in this project was tailored to Port-au-Prince and based on known challenges in the area and considered available local resources and needs. We trust the ability to follow up the microplanning exercises with modified program strategies, such as developing new social mobilization tools or distribution methods, in response to the results of the exercise was essential. However, this requires sufficient time is available between the microplanning exercise and the MDA in order to implement these changes.

The exercise also highlighted the importance of programs’ challenging their assumptions about the reasons for low MDA coverage. The microplanning strategy uncovered barriers to MDA compliance that were unexpected and previously unknown challenges. This allowed stakeholders to leverage local knowledge to identify solutions that were acceptable to MDA volunteers and targeted communities. This is consistent with findings from other studies that have suggested that such grassroots engagement can yield a better understanding of the MDA, lead to a shared management of resources, and responsibilities, and can ultimately lead to higher treatment coverage [13, 42].

This evaluation had some limitations. Since Port-au-Prince is a large and complex urban setting, it was not possible to engage all relevant actors in the microplanning workshops (e.g., private medical sector who are influential on many individuals’ health decisions making and management of gated communities who determine the ability to access these communities). The microplanning evaluation surveys were limited by self-reporting of performance and engagement indicators, and hence, we cannot assert that microplanning led to increases in specific knowledge or performance metrics as microplanning was included as a broader MDA improvement initiative. Thus, it is not possible to attribute improvements in 2018 MDA coverage to any one improvement strategy. However, the increase in MDA coverage in all five communes in 2018 after years of gradual decline and the use of microplanning results in other MDA improvement strategies supports our belief that microplanning played a key role in improving MDA drug coverage in Port-au-Prince in 2018. Coverage was highest in the earliest treating communes and decreased sequentially, which we hypothesize was due to individuals from neighboring communes being treated and highlights the challenges of assessing coverage during post-based MDA in complex urban settings.

We believe the results of this evaluation support that microplanning should be considered in areas that are undergoing MDA for NTDs, particularly areas that have had historic challenges in achieving adequate MDA coverage. Microplanning may be particularly useful in areas where it is challenging to define the boundaries of work areas, and where the program has faced challenges in providing feedback to MDA volunteers and staff. Finally, it is important to note that microplanning is an iterative process and is ideally employed over successive rounds of MDA.

## Data Availability

The authors confirm that the data supporting the findings of this study are available within the article and its supplementary materials.

## Acknowledgments

We would like to give special thanks to the community drug distributors, community promoters, community leaders, and other stakeholders who enthusiastically participated in the microplanning sessions and for their everyday efforts to eliminate LF in Haiti. We would also like to thank our partners, namely the Ministry of Public Health and Population (MSPP) staff, RTI International, the Carter Center, U.S. Agency for International Development (USAID), the Pan-American Health Organization (PAHO), and the University of Notre Dame for their collaborative efforts to improve MDA coverage in Port-au-Prince. We would like to thank Dr. Ryan Wiegand and Dr. Andrew Hill for their assistance in generating the alluvial diagrams.

---

**Supplemental Fig 1.**
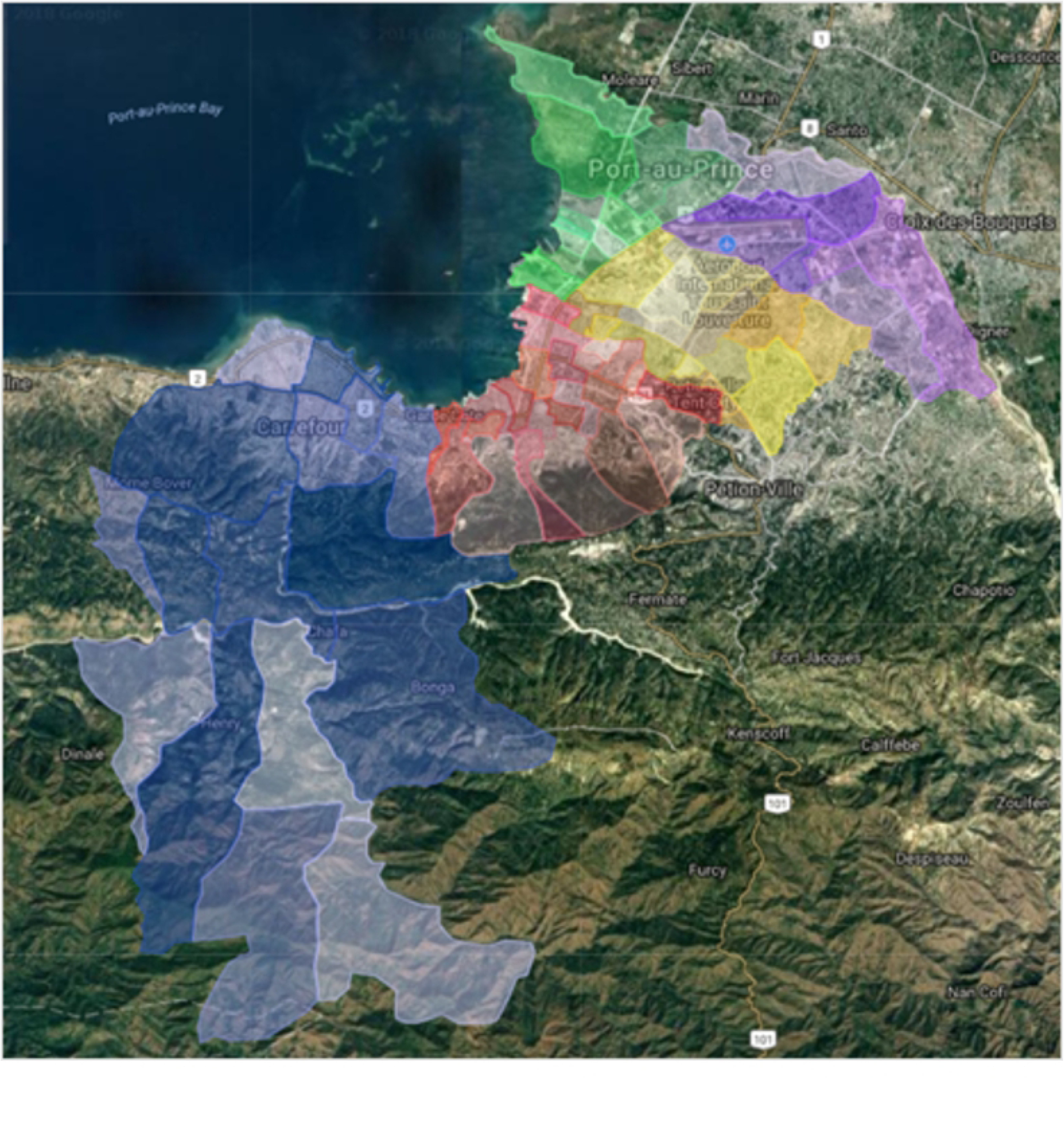
Final supervision area (SA) boundaries for 2018 mass drug administration (MDA) for community leaders (CLs) by commune in Port-au-Prince, Haiti. A map generated by LF program staff in consultation with community leaders during microplanning workshops detailing supervision areas for each community leader for five communes in Port-au-Prince, Haiti. Areas are color coded by commune with shades of blue representing Carrefour, red representing Port-au-Prince, yellow representing Delmas, purple representing Tabarre, and green representing Cité-Soleil.

## References

1. World Health Organization, Global programme to eliminate lymphatic filariasis: progress report, 2017. Weekly epidemiological record, 2018. 93(44): p. 589–604.

2. World Health Organization, Guideline - Alternative mass drug administration regimens to eliminate lymphatic filariasis. 2017: Geneva.

3. Stolk, W.A., et al., Are alternative strategies required to accelerate the global elimination of lymphatic filariasis? Insights from mathematical models. Clin Infect Dis, 2018. 66(suppl_4): p. S260–s266.

4. Babu, B.V. and G.R. Babu, Coverage of, and compliance with, mass drug administration under the programme to eliminate lymphatic filariasis in India: a systematic review. Trans R Soc Trop Med Hyg, 2014. 108(9): p. 538–49.

5. Krentel, A., P.U. Fischer, and G.J. Weil, A review of factors that influence individual compliance with mass drug administration for elimination of lymphatic filariasis. PLoS Negl Trop Dis, 2013. 7(11): p. e2447.

6. Babu, B.V. and S.K. Kar, Coverage, compliance and some operational issues of mass drug administration during the programme to eliminate lymphatic filariasis in Orissa, India. 2004(1360-2276 (Print)).

7. Wynd, S., et al., Qualitative analysis of the impact of a lymphatic filariasis elimination programme using mass drug administration on Misima Island, Papua New Guinea. Filaria Journal, 2007. 6(1): p. 1.

8. Babu, B.V. and S. Mishra, Mass drug administration under the programme to eliminate lymphatic filariasis in Orissa, India: a mixed-methods study to identify factors associated with compliance and non-compliance. Trans R Soc Trop Med Hyg, 2008. 102(12): p. 1207–13.

9. Mitchell, E., et al., Community perceptions and acceptability of mass drug administration for the control of neglected tropical diseases in Asia-Pacific countries: A systematic scoping review of qualitative research. PLoS Negl Trop Dis, 2022. 16(3): p. e0010215.

10. Kisoka, W.J., et al., Factors influencing drug uptake during mass drug administration for control of lymphatic filariasis in rural and urban Tanzania. PLOS ONE, 2014. 9(10): p. e109316.

11. Ames, H.M.R., et al., Community and drug distributor perceptions and experiences of mass drug administration for the elimination of lymphatic filariasis: A rapid review of qualitative research. Adv Parasitol, 2019. 103: p. 117–149.

12. Kisoka, W.J., et al., Community members’ perceptions of mass drug administration of mass drug administration for control of lymphatic filariasis in rural and urban Tanzania. J Biosoc Sci, 2016. 48(1): p. 94–112.

13. Babu, B.V., et al., Use of an inclusive-partnership strategy in urban areas of Orissa, India, to increase compliance in a mass drug administration for the control of lymphatic filariasis. Ann Trop Med Parasitol, 2006. 100(7): p. 621–30.

14. Njomo, D.W., et al., Increasing coverage in mass drug administration for lymphatic filariasis elimination in an urban setting: a study of Malindi Town, Kenya. PLoS One, 2014. 9(1): p. e83413.

15. Krentel, A., et al., Improving coverage and compliance in mass drug administration for the elimination of LF in two ‘endgame’ districts in Indonesia using micronarrative surveys. PLoS Negl Trop Dis, 2016. 10(11): p. e0005027.

16. World Health Organization and Pan American Health Organization, Microplanning manual to guide implementation of preventative chemotherapy to control and eliminate neglected tropical diseases. 2022, World Health Organization and Pan American Health Organization: Geneva.

17. King, M.H., et al., The provision of basic medical care health microplanning: a systems approach to appropriate technology. 1977. 199(1134): p. 61–68.

18. World Health Organization, Reaching Every District (RED): A guide to increasing coverage and equity in all communities in the African Region, 2017 revision, World Health Organization, Editor. 2017, World Health Organization: Brazzaville.

19. Anya, B.-P.M., et al., Contribution of polio eradication initiative to strengthening routine immunization: Lessons learnt in the WHO African region. Vaccine, 2016. 34(43): p. 5187–5192.

20. World Health Organization, Microplanning for immunization service delivery using the Reaching Every District (RED) strategy. 2009, World Health Organization: Geneva.

21. Gidado, S.O., et al., Outreach to underserved communities in northern Nigeria, 2012-2013. J Infect Dis, 2014. 210 Suppl 1: p. S118–24.

22. Dougherty, L., et al., From paper maps to digital maps: enhancing routine immunisation microplanning in Northern Nigeria. BMJ Glob Health, 2019. 4(Suppl 5): p. e001606.

23. Cowan, F.M., et al., Strategies to promote the meaningful involvement of sex workers in HIV prevention and care. Curr Opin HIV AIDS, 2019. 14(5): p. 401–408.

24. Bhattacharjee, P., et al., Micro-planning at scale with key populations in Kenya: Optimising peer educator ratios for programme outreach and HIV/STI service utilisation. PLoS One, 2018. 13(11): p. e0205056.

25. Kamanga, A., et al., Open-source satellite enumeration to map households: planning and targeting indoor residual spraying for malaria. Malar J, 2015. 14: p. 345.

26. Arroz, J.A.H., et al., Implementation strategies to increase access and demand of long-lasting insecticidal nets: a before-and-after study and scale-up process in Mozambique. Malar J, 2017. 16(1): p. 429.

27. Paruthi, R. and P.K. Dutta, Reproductive and child health programme. Indian J Public Health, 2002. 46(3): p. 72–7.

28. Enkhtuya, B., et al., Reaching every district - development and testing of a health micro-planning strategy for reaching difficult to reach populations in Mongolia. Rural Remote Health, 2009. 9(2): p. 1045.

29. Checchi, F., et al., Validity and feasibility of a satellite imagery-based method for rapid estimation of displaced populations. Int J Health Geogr, 2013. 12: p. 4.

30. Ali, D., et al., A cost-effectiveness analysis of traditional and geographic information system-supported microplanning approaches for routine immunization program management in northern Nigeria. Vaccine, 2020. 38(6): p. 1408–1415.

31. Barau, I., et al., Improving polio vaccination coverage in Nigeria through the use of Geographic Information System technology. The Journal of Infectious Diseases, 2014. 210(suppl_1): p. S102–S110.

32. Kamadjeu, R., Tracking the polio virus down the Congo River: a case study on the use of Google Earth in public health planning and mapping. Int J Health Geogr, 2009. 8: p. 4.

33. Lemoine, J.F., et al., Controlling Neglected Tropical Diseases (NTDs) in Haiti: Implementation Strategies and Evidence of Their Success. PLoS Negl Trop Dis, 2016. 10(10): p. e0004954.

34. Streit, T.D., L.; Oscar, R.; Lemoine, J.F.; Purcell, N. C.; Keller, A.; Lammie, P. J.; Deming, M.; Beau De Rochars, V. E. M, Mass drug administration for the elimination of lymphatic filariasis--Port-au-Prince, Haiti, 2011-2012. MMWR Morb Mortal Wkly Rep, 2013. 62(23): p. 466–8.

35. Wodnik, B.K., et al., The roles of stakeholder experience and organizational learning in declining mass drug administration coverage for lymphatic filariasis in Port-au-Prince, Haiti: A case study. PLoS Negl Trop Dis, 2020. 14(5): p. e0008318.

36. Hartung, C., et al., Open data kit: tools to build information services for developing regions, in Proceedings of the 4th ACM/IEEE International Conference on Information and Communication Technologies and Development. 2010, Association for Computing Machinery: London, United Kingdom. p. Article 18.

37. Lam, T.C.M. and P. Bengo, A comparison of three retrospective self-reporting methods of measuring change in instructional practice. The American Journal of Evaluation, 2003. 24(1): p. 65–80.

38. Goedhart, H. and J. Hoogstraten, The retrospective pretest and the role of pretest information in evaluative studies. 1992. 70(3): p. 699–704.

39. Pratt, C.C., W.M. McGuigan, and A.R. Katzev, Measuring program outcomes: Using retrospective pretest methodology. 2000. 21(3): p. 341–349.

40. Howard, G.S., P.R. Dailey, and N.A. Gulanick, The feasibility of informed pretests in attenuating response-shift bias. 1979. 3(4): p. 481–494.

41. Howard, G.S. and P.R. Dailey, Response-shift bias: A source of contamination of self-report measures. Journal of Applied Psychology, 1979. 64(2): p. 144–150.

42. Silumbwe, A., H. Halwindi, and J.M. Zulu, How community engagement strategies shape participation in mass drug administration programmes for lymphatic filariasis: The case of Luangwa District, Zambia. PLoS Negl Trop Dis, 2019. 13(11): p. e0007861.

